# Development of a digital platform for the equitable promotion of mother and child health among marginalized populations: Formative study from a lower-middle income country

**DOI:** 10.1101/2023.04.06.23288277

**Authors:** Zaeem ul Haq, Ayesha Naeem, Durayya Zaeem, Mohina Sohail, Noor ul Ain

## Abstract

**Background:** Health inequities exist between and within countries and communities for maternal and child health, nutrition, and early childhood development. Socially excluded bear the major brunt of this disparity. Innovative ways of providing healthcare are required to meet the needs of such populations. We report the development and feasibility testing of *Sehat Ghar* (Health House), an android-based digital application for volunteer health workers from a population not covered by Primary Healthcare (PHC).

**Methods:** We carried out a mixed-methods study with three steps. First, we conducted 13 in-depth interviews and two Focus Group Discussions (FGDs) with stakeholders to explore the gaps in community knowledge and practices. To address these gaps, we developed the *Sehat Ghar* App, comprised of video-based health education to demonstrate practices that mothers and families need to adopt. Second, we trained ten volunteer Community Health Workers (CHWs) from the same community to deliver health education using the App, and assessed their knowledge and skill improvement. Third, these CHWs visited pregnant and lactating mothers at home, that we monitored using a structured observation list.

**Results:** Initial exploration revealed a need for health-related knowledge and suboptimal healthcare utilization from neighbouring public hospitals. *Sehat Ghar* employed behaviour change techniques, including knowledge transfer, improving mothers’ self-efficacy, and enhanced family involvement in mother and childcare to address this. Volunteer women were trained from the community, who, after the training, showed a significant improvement in mean knowledge score [Before: *M* = 8.00 (*SD* = 1.49), After: *M* = 11.40 (*SD* = 1.43), p=.0007]. Our monitoring found these CHWs excellent in their interaction with mothers and excellent or very good in using the App. The CHW and her community reported their liking and satisfaction with the App and wanted its delivery on a regular basis.

**Conclusions:** The digital application *Sehat Ghar* is a simple, easy-to-use resource for CHWs and is acceptable to the community. Mothers appreciate the content and presentation and are ready to incorporate its messages into their daily practices. The real-world effectiveness of the innovation is currently being tested on 250 mother-infant pairs. With its usefulness and adaptability, and the rapidly spreading mobile phone and Internet technology, the innovation can educate communities at a large scale in a minimum amount of time, contributing to equitable coverage of health services in resource-constrained settings.

## INTRODUCTION

Inequality, disparity, and inequity are commonly discussed in the public health literature. While inequality and disparity are interchangeable and refer to the differences in health determinants and health outcomes of different social and geographical groups, inequity is a subset of this concept [1–3]. According to the World Health Organization (WHO), health inequity is the difference in health status or the distribution of health resources between population groups and arises from the social conditions in which people are born, grow, live, work, and age [4]. Inequity is systematically associated with avoidable, unfair, and unjust elements and is strikingly visible among the rural and slum populations [5,6]

Pakistan is one of the most urbanized countries in South Asia, faced with the challenge of burgeoning slum populations. Currently, 47% of the total urban population lives in the nine biggest cities; 50% are in slums and squatter settlements [7]. Growing urban populations that ultimately expand, enclosing the adjacent villages and smaller towns, along with factors like climate change that cause the migration of unskilled workers from rural to urban areas, are the reasons for upward slum dwellings in the country [7,8]. The informal settlements usually fall outside the mainstream health and governance system, which is already stretched and unable to provide for even the well-settled populations [9,10]. Gender intersectionality further adds to the health challenge for women and children in these areas [11].

It is, therefore, not surprising that the survival and health of mothers and children is an issue with chronic and striking inequality. Of the 287,000 women who die during and following pregnancy and childbirth [12] and 5.3 million children who die before reaching the age of 5 years [13], most belong to low-income settings [14,15]. The maternal mortality ratio in Low to Middle-Income Countries (LMICs) in 2017 was 462 per 100 000 live births, compared to 11 per 100 000 live births in high-income countries [12]. Within-country disparities are also alarming, with disadvantaged populations living in slum areas having much higher mortality [16]. There is an urgent need to address the health requirements of mothers and children living in slum areas.

Biomedical interventions are available to address maternal and child survival and health requirements [17,18]. A significant challenge, however, is the delivery of these interventions to address inequity and improve access of the poor and marginalized populations to the essential set of health services [5]. Information Technology (IT) can help expand the delivery of these services [19–22]. With higher access to mobile phones and the Internet in Pakistan, innovative ways of increasing outreach involving mobile technology can help meet such populations’ needs [23]. However, there can be apprehensions about the availability of the Internet and the feasibility of IT-based services to marginalized communities in slum areas.

The past decade has seen an upsurge in using technology-based interventions. However, these studies focus more on effectiveness, while the “how to” part of these interventions remains unclear [24]. In this paper, we report the development and feasibility testing of *Sehat Ghar* (Health House), an Android-based digital application that Community Health Workers (CHWs) from the local area are using to promote mother and child health to the families living in two of the 34 squatter settlements of Islamabad, Pakistan. The results of this formative study will inform the ongoing trial and help other researchers aiming to use digital technology to empower communities and strengthen primary healthcare systems.

## METHODS

### Aims and objectives

To develop a tailored intervention that fulfills the knowledge and skill-building needs of a community not covered by the primary healthcare system. The specific objectives were as follows:

- Explore the perceptions, practices, and related gaps about mother and child health and child development in the community.
- Design a behaviour change communication program to address these gaps.
- Assess the feasibility of the program on a sub-sample of the study.

### Setting

The study involved two slums of Islamabad, Pakistan’s capital, with a geographical area of 906 Km^2^ and a population of over two million [25]. The city has at least 34 slum areas inhabited by 85,000 people [26]. Many of these slums have poor water, sanitation and hygiene (WASH) conditions and are devoid of public-funded Primary Health Care (PHC) services [27]. We focused our study on France and Rimsha colonies, two squatter settlements located almost in the city’s center, with a population of 8,000 and 10,000, respectively [28]. The residents of both dwellings mainly belong to the Christian faith and labourer class and have low socioeconomic status and poor coverage of PHC services. Water-borne and other environment-related health problems are common; mothers and children suffer the most in these areas [10,27].

### Study design

It was a mixed-methods, participatory research design (Figure 1) in line with our earlier, community-based, formative studies [29]. The study comprised three phases: 1) stakeholder consultations and intervention design, 2) deployment of community health workers, and 3) implementation of the pilot phase.

**Figure 1:**
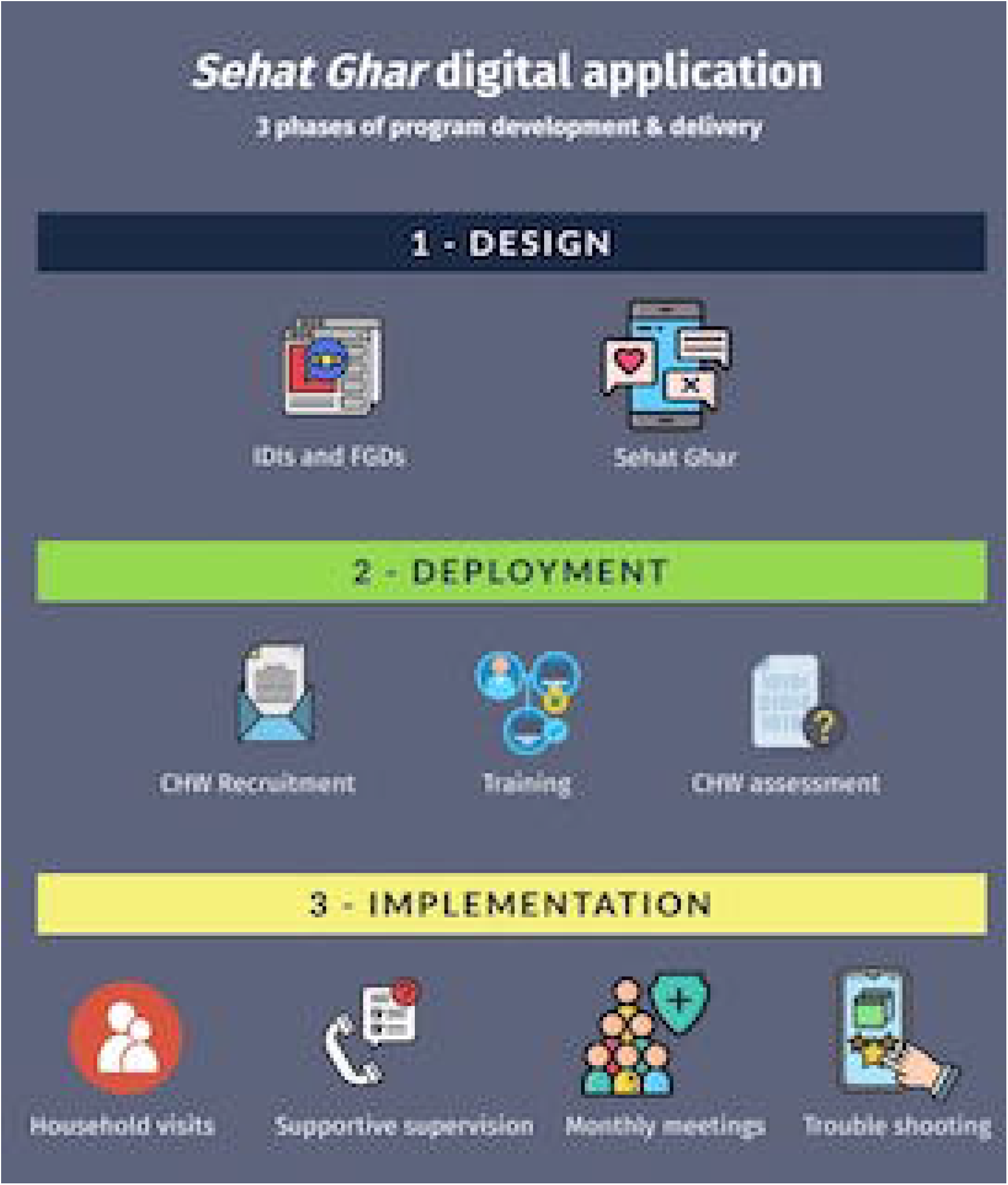
Three phases of *Sehat Ghar* development and implementation.

#### Phase One: Stakeholder participation in the intervention design

This phase involved two undertakings: qualitative explorations to inform our study and development of the intervention.

A) In the absence of functional primary healthcare in the study clusters, building initial contacts in the community was critical. For this, we contacted all Healthcare Providers (HCPs) in the area –formal and informal – and offered them to get involved as a stakeholder in our study. A total of five HCPs (two from Rimsha and three from France colony) agreed. Through them, we identified volunteer women willing to become Community Health Workers (CHW). Female CHWs usually have access to households in their community and educate mothers to improve their birth preparedness [30,31]. Being female, having completed ten years of schooling, living in the same community, and preferably being married were the criteria for their selection. A total of 15 volunteer women were enrolled. These volunteers conducted household visits in their area and enlisted pregnant or nursing mothers from their community willing to participate in our study. We invited ten mothers, five from each cluster, from this list, for in-depth interview (IDI) as part of this formative study.

Overall, we interviewed five HCPs, eight mothers (two did not agree to interview) and two Focus Group Discussions (FGDs) with 15 CHWs during this phase. HCP views about the healthcare-seeking practices of their community during the antenatal, natal, and postnatal periods were explored. In the FGDs with HCWs, we explored the community practices regarding mother and child health, child development, and the gender dimensions prevalent in that culture. Through the IDIs with mothers, we tried to understand the family practices around mother and child nutrition, health-seeking, and child development. Using a semi-structured guide, two team members with extensive qualitative research experience conducted the interviews and FGDs in Urdu, a language convenient to both the community and the study team. The interviews of HCPs and mothers were conducted at the clinic or home, respectively, while the FGDs were conducted in the courtyard of a CHW, who allowed to organize the group at her place. The IDIs took about 30-50 minutes, and a typical FGD was 90 minutes long. All discussions and interviews were tape-recorded, transcribed and translated into English.

B) We used the findings from the qualitative explorations to inform the intervention. The study participants identified knowledge gaps about mother and child health and child development among the mothers and their families. The discussions also revealed among mothers a lack of self-efficacy to organize their actions around better health outcomes for the mother-infant dyad. To address the knowledge gap and provide a platform for the CHW to help the mother and her family through discussions, we developed *Sehat Ghar*—a digital platform to facilitate behaviour change in this population. Applying Human-Centered Design (HCD) principles [32,33], a multidisciplinary team developed this intervention through several stages. To begin with, we summarized the findings from stakeholder consultations in the form of a matrix (Table 1) to highlight the community’s existing knowledge and skills and relevant gaps. With the help of healthcare providers and CHWs, we identified priority behaviours and finalized health education content to address the gaps.

**Table 1:** Gaps in community practices, and the intervention content to address the gaps.

| Themes | Findings | Intervention element |
| --- | --- | --- |
| **Access to primary health care** | No coverage of PHC at the household level<br>Mothers unaware of healthful practices at household level<br>Families tangled in poverty trap | CHW introduced as substitute<br>CHW received training to deliver the intervention<br>Emphasis on behaviors that can be adopted at the household level |
| **Mother's health & nutrition** | Lack of knowledge about antenatal and postnatal care, and practice of getting regular checkups.<br>Lack of knowledge about maternal nutrition<br>Infrequent use of iron tablets<br>Mental stress during pregnancy | Sessions 1-3 on antenatal & postnatal checkups, danger signs and steps to be taken if danger signs appear<br>Emphasis on maternal nutrition, especially iron/folate<br>Family support appreciated and further emphasized especially to husband |
| **Newborn health** | Families opt for formal health facility, yet some also prefer TBA for childbirth<br>Newborns receive checkup only if delivered at health facility<br>Pre-lacteals are common; Breastfeeding delayed for 1-3 days<br>Warm *desi ghee* applied on umbilical cord | Specific information about Skilled Birth Attendant (SBA) and their importance<br>Session 3 on newborn care:<br>Significance of early checkup<br>Identifying and taking action for danger signs in a newborn<br>Early initiation of breastfeeding, delayed bathing for 24 hours<br>Using chlorhexidine for cord care emphasized |
| **Child health & nutrition** | Children taken to hospital only for immunization or in case of emergency<br>Lack of knowledge about duration and significance of exclusive breastfeeding<br>Perception of insufficient milk is common<br>Water and semi solids frequent between 2-5 month<br>Lack of knowledge about child nutrition | Sessions 3-7 focus on child health and encourage families to consult health care provider whenever necessary<br>Session 5 & 6 specifically talk about preventable diseases, their symptoms, and child immunization<br>Sessions 4-7 emphasize exclusive breastfeeding for 6 months, and explain child's nutritional requirements with growing age<br>These sessions also include homemade recipes for adding semi-solid diets for children above 6 months. |
| **Environment for child's development** | No awareness and understanding about child stimulation and play<br>Families focus on child nutrition, and link nutrition only with physical growth | Session 4-7 explain stimulation and play and also provide age-appropriate play activities for children<br>Content on making toys from everyday items to enhance child stimulation and play |

Our key informant mothers and CHWs preferred stories or dramas to convey health information. We engaged a creative team comprised of a graphic designer, a scriptwriter, and a film producer to develop the storyline and record short docudramas to address the information gaps. Once the graphics and audio-visual clips were ready, we collaborated with the app development team to transfer the entire set into a user-friendly, easy-to-navigate digital package. Called *Sehat Ghar*, this digital application was installed on a tablet for the CHWs to use in their counselling sessions with the mothers. Upon CHW’s recommendation, a wall calendar for mothers was also developed that displayed the same graphics and short messages as were in the video. This calendar would remind the mother about the actions she needed to carry out until the CHW’s next visit. Input from CHWs and the community was incorporated into these pieces at all stages of the intervention development.

#### Phase Two: Training and Deployment of Community Health Workers

We offered a two-day training to all 15 CHWs who enrolled in our study. Due to personal or family reasons, five dropped at different time points and a final set of 10 CHWs completed all steps of training and apprenticeship. The entire process comprised a two-day training followed by one week of apprentice work in the field and one day’s refresher training after the CHW completed their fieldwork. The training curriculum comprised video-based health education content to address a) Gaps in the knowledge about mother and child health, b) Interpersonal communication, and c) Skill development for using the tablet and the *Sehat Ghar* app. We assessed CHWs’ knowledge through a questionnaire before and after the training and obtained their feedback qualitatively once they started their household visits.

#### Phase Three: Implementation

Once the CHW started their home visits, a field supervisor who was also involved in CHW training regularly visited them to observe their real-world performance and practice. Using a 21-item (yes/no) checklist especially developed for this study, she recorded observations on *fidelity to intervention* (6 items), *mastery of using the tablet* (5 items), and *Command over the content* (10 items). At the end of each visit, the supervisor appreciated a satisfactory CHW performance and discussed any areas of improvement. Data from the initial 20 visits (2 visits per CHW) was analyzed to carry out the performance ranking of CHWs and draw lessons for collective feedback to the CHWs. On the first follow-up after three days, qualitative feedback was also obtained from CHWs about the *Sehat Ghar* application and tablet use. In addition, the supervisor also obtained qualitative feedback from mothers about the usefulness of CHW visits and the content of *the Sehat Ghar* application.

### Data management and analysis

For qualitative data, the team transcribed the Urdu discussions from interviews and FGDs and translated these into English. Two team members independently read and coded the initial five transcripts to develop a code list. This initial code list was discussed to see congruence between the two coders and divergent points resolved for a single consensus code sheet. The analysis identified significant statements across all transcripts using inductive techniques [34,35]. These meaning statements were clustered together to identify themes. We gave weightage to recurring themes; however, we also paid attention to the divergent themes or points that were not shared by many participants but appeared significant [36]. For quantitative analysis, we compared mean scores from the knowledge test of CHWs using a paired, one-tailed t-test, as the data at both time points came from the same set of participants for the hypothesized increase in CHW knowledge after the training. The quantitative data during implementation monitoring were rank-ordered to make supportive supervision decisions during the implementation.

### Ethical considerations

The study team consisted of medical, public health and sociology background researchers, having formal training in mixed-methods research and significant practice of formative and process evaluations. All team members had the experience of collaborating with communities in the design and delivery of health interventions. Most of the team members were female, as the nature of work required close collaboration with mothers and other female members of their households. At any stage, the team did not offer any incentives or payments to the ultimate beneficiary participants in this research. The Institutional Review Board of the Health Services Academy, Islamabad, Pakistan, approved the study. Grand Challenges Canada funded this study through a seed grant (ST-POC-1808-17445), and this formative phase was completed from April to August 2018.

## RESULTS

### Phase one: Stakeholder consultations and intervention design

The following five themes emerged from the IDIs with health providers and mothers and the FGDs with community health workers.

1. *Access to primary healthcare is poor:* In a community with low literacy among women and no outreach service as part of PHC, women relied on untrained, traditional birth attendants for an antenatal checkup (ANC) and advice. Very few went to public hospitals for their routine ANC, even though the hospitals were not far. For childbirth, however, they usually went to the public hospital and faced hardships because of not having prior registration in the hospital system. Postnatal checkups were minimal; most respondents thought their poverty was a barrier to good nutrition, healthcare, and education.
2. *Inadequate knowledge about mother’s health and nutrition:* Most women were unaware of their physical and nutritional requirements during pregnancy and after childbirth. Similarly, they did not know when to have a checkup during pregnancy and after childbirth and how many times a child needs to get vaccinated. Those who sought healthcare from the formal sector needed more empowerment to ask questions about their health and medication. Information was scant about the relationship between nutritious food and minerals (e.g., iron) with the development of a baby’s brain in the mother’s womb.
3. *Inadequate attention to newborn care:* A newborn’s only checkup was immediately after birth. Afterwards, the baby would see a health provider during immunization or when sick. While the WHO advises delaying the first bath for 24 hours, families practiced an average delay of five days to the first bath during winters and, as per the situation, in summers. Applying warm homemade butter (desi ghee) on the umbilical cord to keep it healthy was a common practice. Breastfeeding initiated with a delay of up to three days as mothers thought that milk letdown starts around the third day after childbirth.
4. *Lack of knowledge about child health and nutrition:* The importance of adequate breastfeeding and complementary feeding was a distant concept. Mothers stopped exclusive breastfeeding earlier than six months because of the perception that the mother’s milk alone does not fulfill the child’s nutritional needs. Some mothers who intended to breastfeed for two years stopped the practice when they became pregnant again. The family would start adding different liquid and semi-solid foods to the infant’s diet and continue the item the child accepted according to their perception.
5. *The home environment offers mixed conditions for child development:* Generally, the husband and other family members share the household chores with the expecting mother, which could positively reflect a child’s development. However, a child’s development was an alien concept to these families. Most women described development in terms of physical growth, and very few recognized “play” as part of a child’s development. On probing, the factors like poverty, mental stress, lack of space for play within and outside the house, and the overarching fears about the mother’s health and baby’s survival appeared as strong barriers.

### *Sehat Ghar*, the intervention

To address the lack of access to PHC in these communities and to help improve their knowledge, skill, and self-efficacy, we designed the *Sehat Ghar* intervention comprised of seven sessions that used 14 live-action videos and a health calendar (Table 1). Delivered by the CHW at the mother’s house at monthly intervals, the sessions start from the last trimester of pregnancy until the child becomes six-month-old. The content delivers timely and appropriate messages on maternal health & nutrition, child health & nutrition, the importance of skilled birth attendance, immunization, and a positive home environment.

The sessions include two action videos for discussion during CHW’s visit. The two videos present a contrasting story through characters played by human actors. The first story narrates a household where the family environment does not encourage a mother’s healthful nutrition, and the mother and her baby ultimately face adverse health consequences. In the same session, the second video shows a family with a similar socioeconomic status but positive thinking and self-belief, dealing with economic problems yet ensuring healthy food for the mother and the baby. After showing the videos, the CHW discusses with the mother and her family similar problems they may be facing and the best ways to jointly solve them, as the positively deviant family [37] was doing in the video. Using the simplified cognitive behavioural training technique used in community-based studies earlier [29], the CHW finally discusses the practical activities that a mother and family could implement to solve those problems.

The mode of delivering this content is interactive and non-didactic. The CHW emphasizes interspousal discussion for problem-solving by making the best use of the available household resources. The CHW engages the mother-in-law more in a family where the husband is not ready to participate in household discussions. Homemade toys and low-cost nutritional solutions are proposed when a family has financial constraints. We theorized (Figure 2) that CHW visits to deliver *Sehat Ghar* content will improve the knowledge and self-efficacy of mothers about the steps that can be ensured at home and support family practices, including facilitating the mother to go to a nearby public health facility for healthcare. The combined result of these practices will be an improvement in the mother’s and child’s health and development.

**Figure 2:**
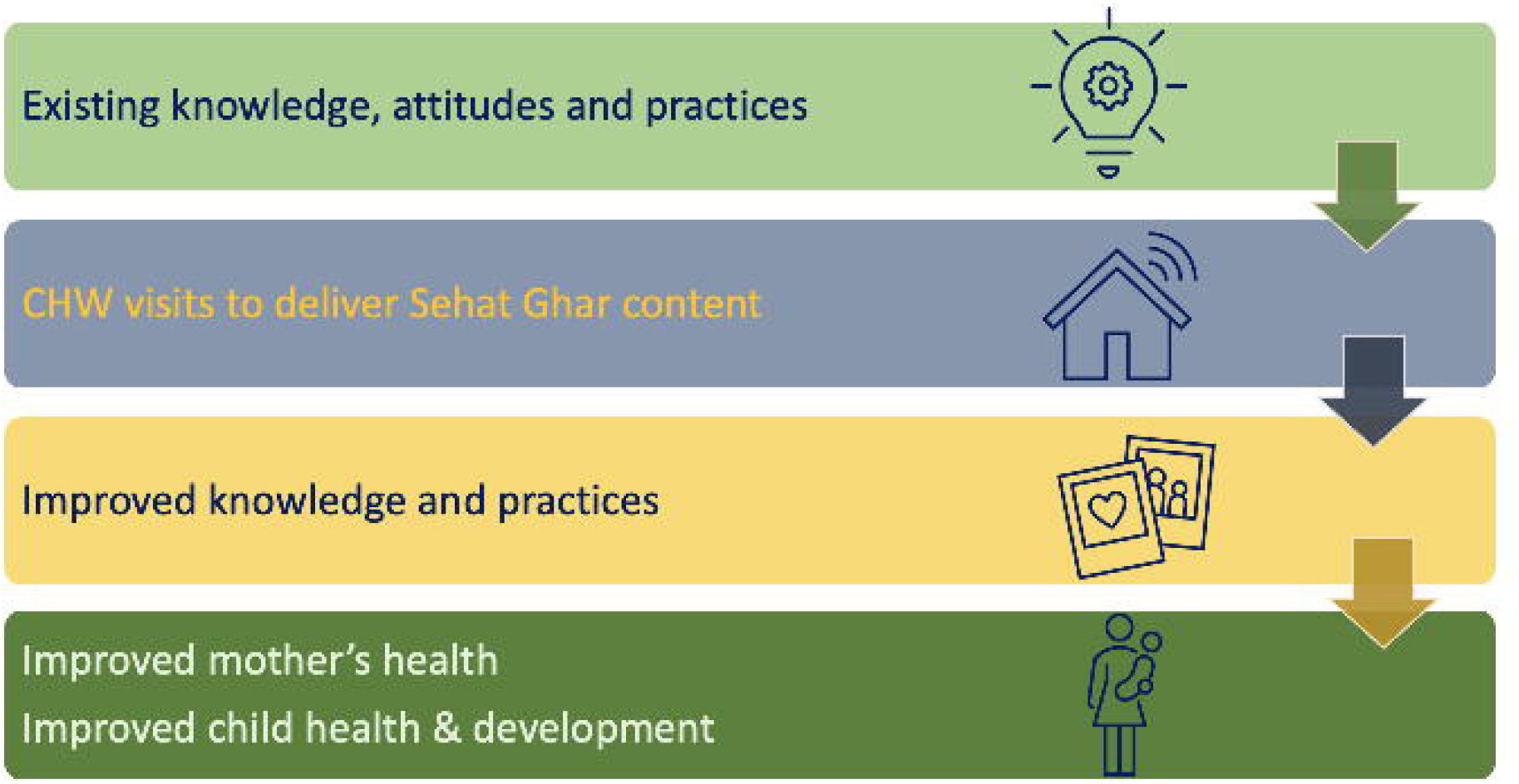
Pathway of change initiated by *Sehat Ghar* intervention.

### Phase two: CHW training and deployment

Out of the 15 potential CHWs, two opted out because they could not spare time from home chores, while three needed to get permission from their elders. We worked with a final cohort of 10 CHWs, all female, having minimal schooling of 10 years and belonging to the same community. The assessment of knowledge of these CHWs before and after the training (Table 2) showed significant improvement (Mean knowledge score before training *M* = 8.00 (*SD* = 1.49), after-training *M* = 11.40 (*SD* = 1.43), p=.0007) after the training.

**Table 2:**
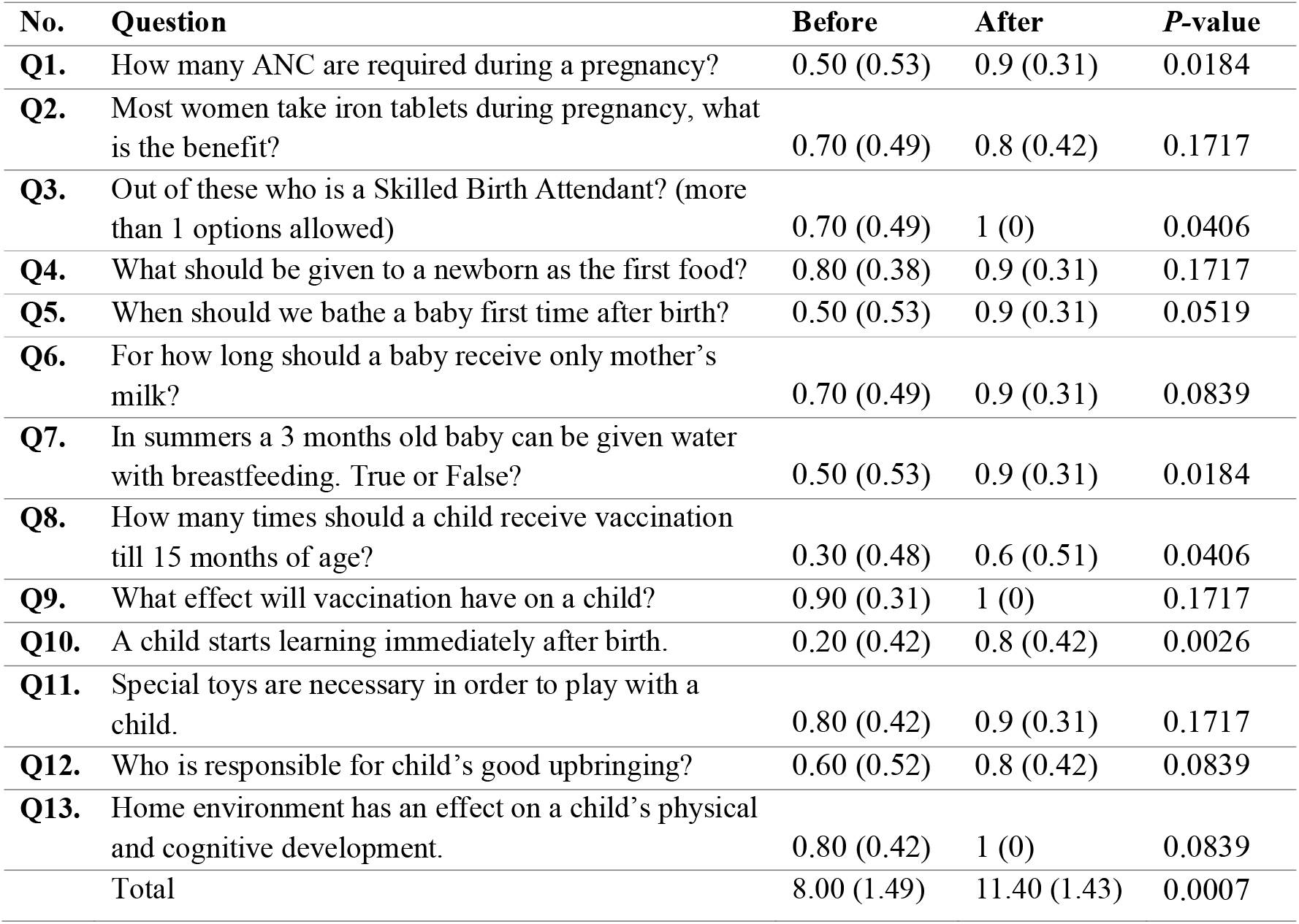
Mean knowledge scores of CHW before and after the training (n=10)

| **No.** | **Question** | **Before** | **After** | **P-value** |
| --- | --- | --- | --- | --- |
| **Q1.** | How many ANC are required during a pregnancy? | 0.50 (0.53) | 0.9 (0.31) | 0.0184 |
| **Q2.** | Most women take iron tablets during pregnancy, what is the benefit? | 0.70 (0.49) | 0.8 (0.42) | 0.1717 |
| **Q3.** | Out of these who is a Skilled Birth Attendant? (more than 1 options allowed) | 0.70 (0.49) | 1 (0) | 0.0406 |
| **Q4.** | What should be given to a newborn as the first food? | 0.80 (0.38) | 0.9 (0.31) | 0.1717 |
| **Q5.** | When should we bathe a baby first time after birth? | 0.50 (0.53) | 0.9 (0.31) | 0.0519 |
| **Q6.** | For how long should a baby receive only mother's milk? | 0.70 (0.49) | 0.9 (0.31) | 0.0839 |
| **Q7.** | In summers a 3 months old baby can be given water with breastfeeding. True or False? | 0.50 (0.53) | 0.9 (0.31) | 0.0184 |
| **Q8.** | How many times should a child receive vaccination till 15 months of age? | 0.30 (0.48) | 0.6 (0.51) | 0.0406 |
| **Q9.** | What effect will vaccination have on a child? | 0.90 (0.31) | 1 (0) | 0.1717 |
| **Q10.** | A child starts learning immediately after birth. | 0.20 (0.42) | 0.8 (0.42) | 0.0026 |
| **Q11.** | Special toys are necessary in order to play with a child. | 0.80 (0.42) | 0.9 (0.31) | 0.1717 |
| **Q12.** | Who is responsible for child's good upbringing? | 0.60 (0.52) | 0.8 (0.42) | 0.0839 |
| **Q13.** | Home environment has an effect on a child's physical and cognitive development. | 0.80 (0.42) | 1 (0) | 0.0839 |
|  | Total | 8.00 (1.49) | 11.40 (1.43) | 0.0007 |

In the qualitative feedback after training and during fieldwork, CHWs showed confidence in their competency and community acceptance. They appreciated *Sehat Ghar*, especially its videos. One shared, *“As soon as we play the video, all the training gets refreshed in our mind. Also, the videos give us words to communicate better. We get much support through these videos*.*”* Some CHWs experienced problems because their clients found these videos lengthy and time-consuming—the CHWs improvised by narrating a brief story and going on to the discussion. One CHW shared, *“There are women who do not seem to have enough time. They are always busy with their children or other household chores and make excuses when we visit them. I do not show full videos in that case but tell them the main story. Eventually, I think, these women will develop interest*.*”*

The study team and the CHWs also faced a few difficulties. The absence of a public sector healthcare system for these communities made accessing the community and households challenging. We engaged 15 health workers but worked with ten from the start till the end of the study. There were interruptions to the Internet in field areas, because of which updates to the App could not be instantly downloaded. This connectivity issue was solved by timing all the updates with monthly meetings of the CHWs, which were held at the principal office of the study team, where high-speed Internet was available to facilitate those downloads. The length of the videos was a challenge for some mothers because of needing more time to handle the number of household chores. However, the majority were interested in the characters and stories presented in the videos.

### Phase three: Implementation Monitoring

During household visits observed for 20 families (Table 3), the CHWs showed *fidelity to intervention*. They fulfilled (in more than 90% of visits) all the required steps except for praising the mother and family on accomplishments which were fulfilled during 58% of visits. The *mastery of a tablet* was very good to excellent, as all items received a high score (83%-100%) in all the visits. Five items in *the Completeness of Intervention* received high (85%-92%), while the other five received low to moderate scores (33%-80%) during these visits. The items that did not receive an adequate score included using the main picture to build discussion, giving the mother enough time to speak, using the full session content, including videos, and reminding the mother about the practical tasks.

**Table 3:**
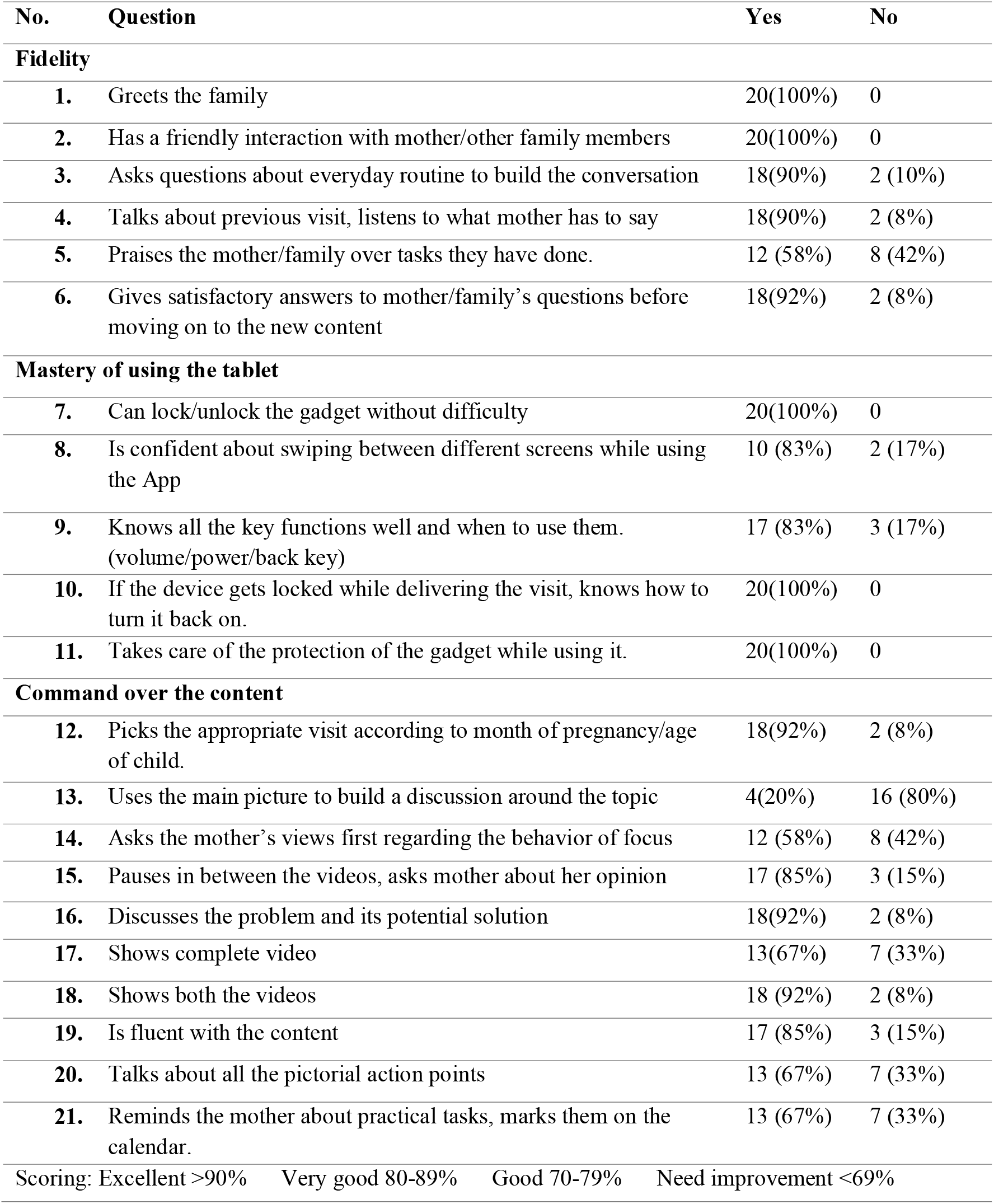
CHW’s performance in engaging with family, and using the tablet and the App.

| No. | Question | Yes | No |
| --- | --- | --- | --- |
| **Fidelity** |  |  |  |
| 1. | Greets the family | 20(100%) | 0 |
| 2. | Has a friendly interaction with mother/other family members | 20(100%) | 0 |
| 3. | Asks questions about everyday routine to build the conversation | 18(90%) | 2 (10%) |
| 4. | Talks about previous visit, listens to what mother has to say | 18(90%) | 2 (8%) |
| 5. | Praises the mother/family over tasks they have done. | 12 (58%) | 8 (42%) |
| 6. | Gives satisfactory answers to mother/family's questions before moving on to the new content | 18(92%) | 2 (8%) |
| **Mastery of using the tablet** |  |  |  |
| 7. | Can lock/unlock the gadget without difficulty | 20(100%) | 0 |
| 8. | Is confident about swiping between different screens while using the App | 10 (83%) | 2 (17%) |
| 9. | Knows all the key functions well and when to use them. (volume/power/back key) | 17 (83%) | 3 (17%) |
| 10. | If the device gets locked while delivering the visit, knows how to turn it back on. | 20(100%) | 0 |
| 11. | Takes care of the protection of the gadget while using it. | 20(100%) | 0 |
| **Command over the content** |  |  |  |
| 12. | Picks the appropriate visit according to month of pregnancy/age of child. | 18(92%) | 2 (8%) |
| 13. | Uses the main picture to build a discussion around the topic | 4(20%) | 16 (80%) |
| 14. | Asks the mother's views first regarding the behavior of focus | 12 (58%) | 8 (42%) |
| 15. | Pauses in between the videos, asks mother about her opinion | 17 (85%) | 3 (15%) |
| 16. | Discusses the problem and its potential solution | 18(92%) | 2 (8%) |
| 17. | Shows complete video | 13(67%) | 7 (33%) |
| 18. | Shows both the videos | 18 (92%) | 2 (8%) |
| 19. | Is fluent with the content | 17 (85%) | 3 (15%) |
| 20. | Talks about all the pictorial action points | 13 (67%) | 7 (33%) |
| 21. | Reminds the mother about practical tasks, marks them on the calendar. | 13 (67%) | 7 (33%) |
| Scoring: Excellent >90% Very good 80-89% Good 70-79% Need improvement <69% |  |  |  |

On average, the CHWs spent 45-60 minutes during a typical household visit. In the qualitative feedback, mothers and their families were happy to have the CHW visit them and educate them about their and the baby’s health. *“Nobody has ever come to visit us as these women do, and no one even comes to give us information verbally*,” said one mother. Mothers appreciated the idea of conveying helpful information through videos and wanted to receive it continuously. They said it increased their knowledge and changed some of their practices. One shared, “*There were many things we did not know or were doing wrong. I was taking iron tablets with milk and learned the correct method through these visits, and now I take them with water*.*”*

## DISCUSSION

This formative study shows that technology-based innovations can be developed to promote mother and child health and child development among families living in slum areas that have no or minimal access to PHC. It is particularly relevant to settings where women have nominal education, families are resource-poor, and deeply embedded cultural practices impede the adoption of healthful behaviours. In a community without the coverage of PHC, this study identified and trained health volunteers who learned using a video-based digital app and then provided health education to mothers and families to facilitate a positive change in their health behaviours.

Involving communities to identify problems and jointly develop solutions is fundamental in areas where PHC coverage does not exist. Through stakeholder consultations as part of this study, we identified three sets of problems that our intervention addressed. First were the knowledge gaps that required providing information to a mother in a way she could understand and remember. The second was a mother’s lack of capacity to recognize why a problem was a problem, which the CHW helped along with jointly working on a solution to that problem. The third was the lack of self-efficacy, without which the perceived threat of a health hazard and the perceived benefit of a suggested action do not translate into practice [38]. Based on our earlier work using cognitive behavioural training [29], we aimed at enhancing self-efficacy by emphasizing activities that a mother could perform, e.g., availing an antenatal checkup or talking to her husband about child spacing. These actions, which initially seemed monumental, became easy and eventually aligned with the mother’s schedule.

In addition to identifying problems in partnership with stakeholders, there were two critical elements to developing this intervention. One the adoption of design principles [32,33,39] to ensure the engagement of stakeholders through all phases of the development and deployment of the intervention. A continuous feedback loop during these phases ensured our design was genuinely human-centred. Paraphrasing Steve Jobs [40], the design was not just the look and feel of the graphics and text; the design was how it all worked according to the preference of our community. Two, collaborating with a multi-talented, creative team in conceptualizing a content that was engaging, memorable, and action oriented. *Sehat Ghar* videos – the output of this collaboration – proved a powerful tool in this regard. The CHWs felt the videos made their job interesting and easy and brought power to the message, the messenger and the program, as has been reported in the past [41].

Some limitations are worth mentioning for a better interpretation of our findings. We wanted to conduct as many interviews and FGDs as required to reach a theoretical saturation point but had to limit our sample given the access issues. In the end, however, we managed a reasonable sample for the qualitative explorations. Secondly, being a research study for a small population, arranging electronic tablets was feasible, which may bring cost issues for the large-scale implementation. However, our App is adaptable to a mobile phone, which can offset these cost implications. Lastly, high-speed Internet may not be available to CHWs in the field for downloading updates. In our context, we integrated updates with the monthly visits of the CHW to the study office, which can be adopted elsewhere.

Technology-based interventions like text-message reminders via mobile phones, robocalls, and social media have been used in the past; their top-down communication and delivery from a distance limit their effectiveness [42,43]. Studies assessing other behaviour change strategies, like messaging campaigns through mass media, have found that they may not be effective when used in isolation [44]. Combining media and technology with face-to-face communication works better [45]. Bringing in health workers, providing training and upgrading their knowledge and skills using phone apps has also received attention [46], just like we did in this study.

However, health content delivered through face-to-face communication by workers trained in cascade setting dilutes the intervention with each step of trickle-down and must be addressed by a “cascade-plus” approach [47]. We embedded our digital App in a cascade-plus training of CHWs in which they were provided more than one job aid and multiple learning opportunities to discuss and improve their skills. One of the reported weaknesses of digital interventions is the need for a behavioural theory upon which the study is modelled [48]. *Sehat Ghar* combined video stories with powerful theoretical concepts of learning from positively deviant families and enhancing self-efficacy for its conceptual framework and multi-stage evaluation [38,49].

The rapid identification of CHWs and quick training on using a digital app can be effective for populations struck by emergencies—a phenomenon that is increasingly becoming common. For example, the torrential rains and floods of 2022 in Pakistan resulted in 33 million people getting affected. About 650,000 among them were pregnant women, out of which 73,000 were likely to give birth in the weeks immediately after these floods [50]. In addition to those giving birth to babies, women and girls seeking access to contraception and other reproductive health treatments also faced challenges [51]. Similarly, the populations affected by wars and conflict, living in camps without organized healthcare, can be served by similar interventions.

Based on the results from this pilot phase, we conclude that developing easy-to-use digital applications like *Sehat Ghar* is feasible and acceptable to the CHWs and the community. Its systematic rollout can contribute to improving health behaviours, including healthcare utilization. Mothers appreciate interesting content like video stories and are willing to incorporate the information into their daily practices. Applying participatory approaches during the formative phase of such interventions is critical for their adoption. The real-world effectiveness of this innovation is being tested in a sample of 250 mother-infant pairs from the same area. With the rapidly spreading Internet technology, such innovations can be a step towards equitable coverage of health services in resource-constrained settings.

## Data Availability

All data produced in the present study are available upon reasonable request to the authors

## List of Abbreviations

ANC: Antenatal Checkup
CHW: Community Health Worker
HCD: Human Centered Design
IT: Information Technology
FGD: Focus Group Discussion
LHW: Lady Health Worker
LMIC: Low to Middle Income Country
PHC: Primary Health Care
WASH: Water, Sanitation, Hygiene
WHO: World Health Organization

## Author Contributions

ZH conceptualized the study. ZH, AN and NA oversaw the data collection and analysis. DZ and MS helped in final analysis and writing different sections. All authors read and approved the final draft.

## Ethics declarations

We obtained ethical approval from the Institutional Review Board of the Health Services Academy.

## Consent for publication

We obtained consent for publication from all study participants.

